# Deep Learning Improves Pre-Surgical White Matter Visualization in Glioma Patients

**DOI:** 10.1101/2020.06.18.20133108

**Authors:** Sandip S Panesar, Vishwesh Nath, Sudhir K Pathak, Walter Schneider, Bennett A. Landman, Michael Iv, Diana Anthony, Tatiana Jansen, Kumar Abhinav, Juan Fernandez-Miranda

**Affiliations:** Department of Neurosurgery, Stanford University, Palo Alto, California, U.S.A.; Electrical Engineering and Computer Science, Vanderbilt University, Nashville, Tennessee, U.S.A.; Learning Research and Development Center, University of Pittsburgh, Pittsburgh, Pennsylvania, U.S.A.; Department of Radiology, Stanford University, Palo Alto, U.S.A.; Biomedical Engineering, Vanderbilt University, Nashville, Tennessee, U.S.A; Department of Neurosurgery, North Bristol NHS Trust, Bristol, United Kingdom

**Keywords:** Deep learning, artificial intelligence, machine learning, tractography, neuro-oncology, glioma, neuroimaging

## Abstract

**Background:** Diffusion tensor imaging (DTI) is a commonly utilized pre-surgical tractography technique. Despite widespread use, DTI suffers from several critical limitations. These include an inability to replicate crossing fibers and a low angular-resolution, affecting quality of results. More advanced, non-tensor methods have been devised to address DTI’s shortcomings, but they remain clinically underutilized due to lack of awareness, logistical and cost factors.

**Objective:** Nath et al. (2020) described a method of transforming DTI data into non-tensor high-resolution data, suitable for tractography, using a deep learning technique. This study aims to apply this technique to real-life tumor cases.

**Methods:** The deep learning model utilizes a residual convolutional neural network architecture to yield a spherical harmonic representation of the diffusion-weighted MR signal. The model was trained using normal subject data. DTI data from clinical cases were utilized for testing: Subject 1 had a right-sided anaplastic oligodendroglioma. Subject 2 had a right-sided glioblastoma. We conducted deterministic fiber tractography on both the DTI data and the post-processed deep learning algorithm datasets.

**Results:** Generally, all tracts generated using the deep learning algorithm dataset were qualitatively and quantitatively (in terms of tract volume) superior than those created with DTI data. This was true for both test cases.

**Conclusions:** We successfully utilized a deep learning technique to convert standard DTI data into data capable of high-angular resolution tractography. This method dispenses with specialized hardware or dedicated acquisition protocols. It presents an economical and logistically feasible method for increasing access to high definition tractography imaging clinically.

## Introduction

Common pre-surgical tractography techniques employ single-shell diffusion tensor imaging (DTI) acquisition protocols ^1^. DTI is known to possess several critical limitations, however, potentially affecting the quality of tractography results ^2–4^. Its major limitation arises from an inability to trace crossing fibers within the voxel, resulting in tracts that appear thin, possess false continuities, or terminate abruptly, relative to “real” anatomy ^5–7^. Alternatives to DTI have been devised to address the crossing-fiber issue and improve angular resolution, including multi-tissue constrained spherical deconvolution (MT-CST) ^8^, high-angular resolution diffusion imaging (HARDI) ^9^ and generalized q-sampling imaging (GQI) ^10^, among others. These techniques generally utilize multi-shell diffusion-weighted MRI acquisitions and yield multi-directional orientation units such as the fiber orientation distribution function (fODF) ^11,12^ or spin distribution function (SDF) ^10^. At present, the majority of these non-tensor sequences are utilized for research purposes, rather than in clinical settings ^6,13^. Relative to DTI, however, acquisition of multi-shell, non-tensor sequences is more expensive and time-consuming ^14^, making them impractical to implement or utilize clinically.

Deep learning is a powerful tool that permits learning of non-linear mappings between a set of inputs and outputs. Nath et al. (2020) presented a deep learning-based pipeline for recovery of MT-CSD from single-shell diffusion data using a residual convolutional neural network (ResCNN) ^14^. In this study, we adapted the deep learning model, trained using normal subjects from the Human Connectome Project (HCP) ^15^, and utilized it to process single-shell DTI data from 2 real tumor cases. We then performed tractography on the output data and compared it qualitatively and quantitatively with tractography conducted using the standard DTI data.

## Materials and Methods

### Subjects

Both subjects gave their consent for their anonymized data to be utilized in this study, which was approved by the Institutional Review Board at **[REDACTED FOR REVIEW]**.

### Subject 1 – Right-sided parietal tumor

A male patient in their 50’s presented with a large right-sided parietal mass of 47 × 35 millimeters, associated with a 1cm midline shift and vasogenic edema. He underwent right-sided craniotomy and tumor resection. This was later determined to be a World Health Organization (WHO) Grade III anaplastic oligodendroglioma **(Figure 1A, B)**.

**Figure 1.**
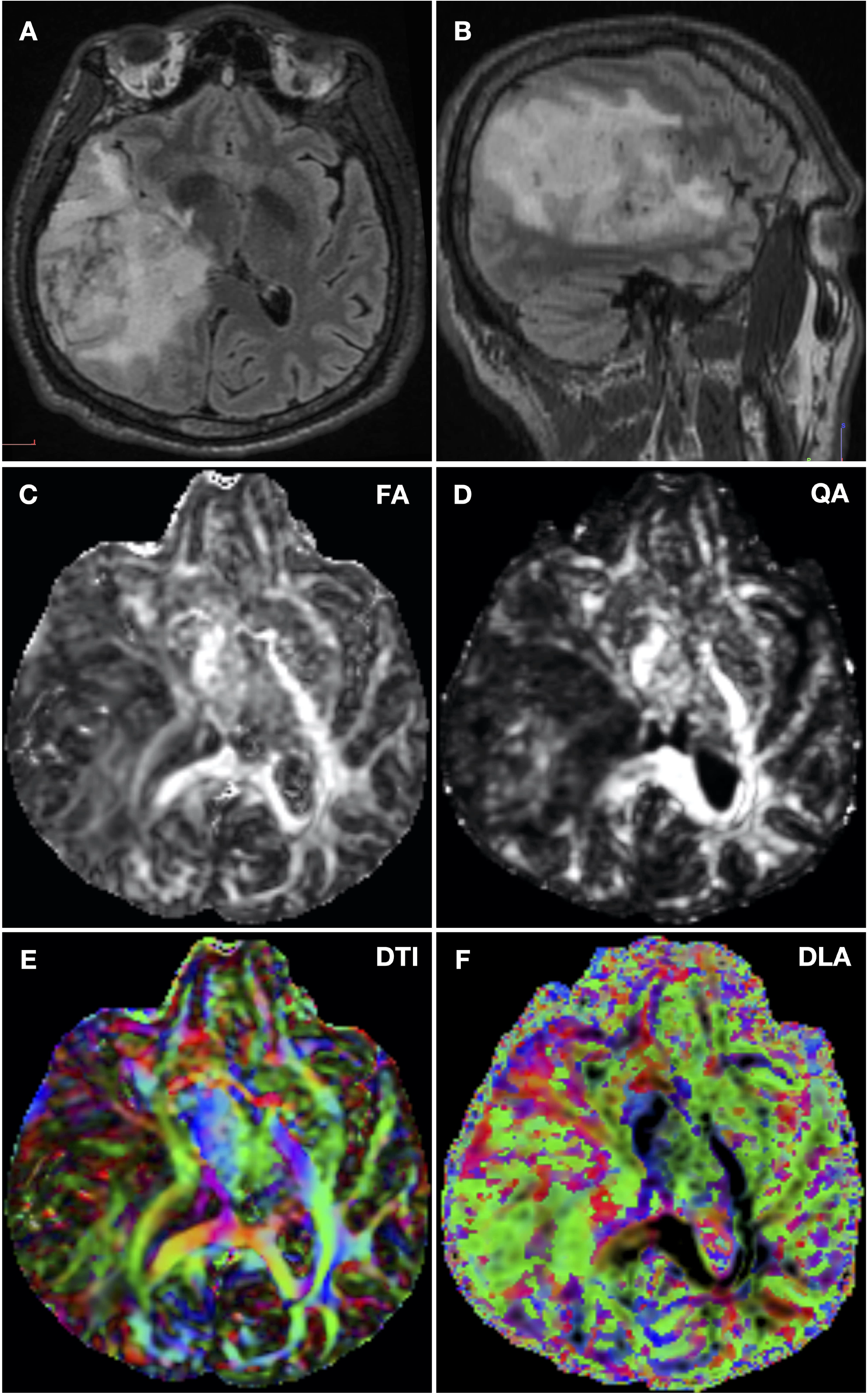
**A –** Axial FLAIR sequence demonstrating large right sided parietal mass in subject 1. The tumor is surrounded with substantial edema, and is causing a degree of midline shift. **B –** Sagittal FLAIR sequence demonstrating the parietal lobe of subject 1. Edema extends into the upper parietal lobe, the occipital lobe and the upper temporal lobe. **C –** Fractional anisotropy (FA) map of subject 1, visible is a loss of white matter architectural integrity in area of the tumor. **D –** Quantitative anisotropy (QA) map of subject 1. The tumor region appears darker compared to the corresponding FA map. **E –** Color diffusion tensor map produced by diffusion tensor imaging (DTI) sequence. Green – anteroposteriorly travelling fibers; Blue-superoinferiorly travelling fibers; Red – laterally travelling fibers. **F –** Color orientation distribution function map produced by deep learning algorithm (DLA) processing demonstrating. Green – anteroposteriorly travelling fibers; Blue-superoinferiorly travelling fibers; Red – laterally travelling fibers.

### Subject 2 – Right-sided temporal tumor

A female patient in their 50’s presented with a right-sided temporal mass of 29 × 20 millimeters. She subsequently underwent a right-sided craniotomy and tumor resection. This was later determined to be a WHO Grade IV glioblastoma **(Figure 2A, B)**.

**Figure 2.**
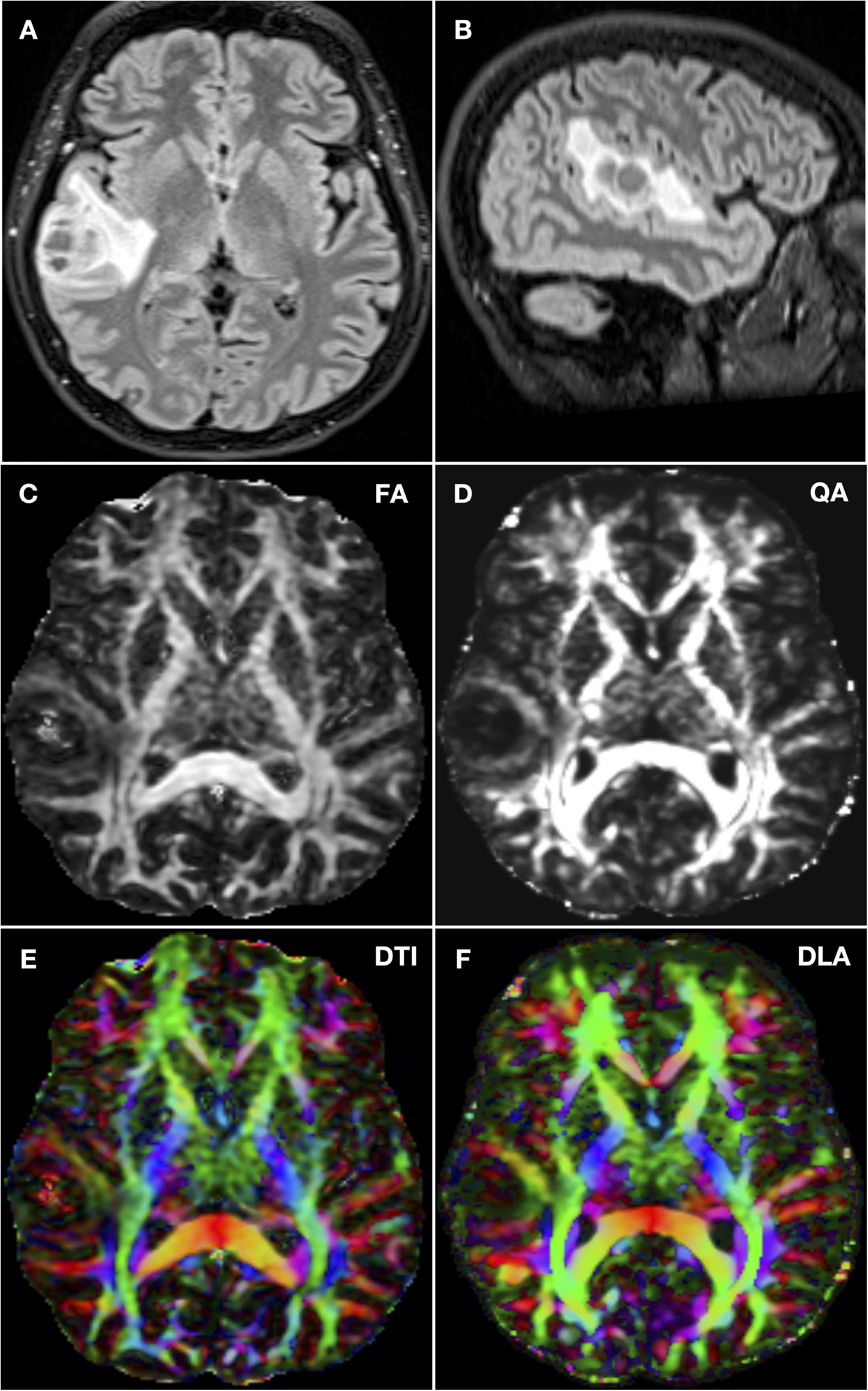
**A –** Axial FLAIR sequence demonstrating right sided parietal mass in subject 2. The tumor is surrounded with some edema. **B –** Sagittal FLAIR sequence demonstrating the superior temporal lobe of subject 2. Edema is distributed longitudinally along this gyrus. **C –** Fractional anisotropy (FA) map of subject 2. **D –** Quantitative anisotropy (QA) map of subject 2. **E –** Color diffusion tensor map produced by diffusion tensor imaging (DTI) sequence. Green – anteroposteriorly travelling fibers; Blue-superoinferiorly travelling fibers; Red – laterally travelling fibers. **F –** Color orientation distribution function map produced by deep learning algorithm (DLA) processing demonstrating. Green – anteroposteriorly travelling fibers; Blue-superoinferiorly travelling fibers; Red – laterally travelling fibers.

### Pre-operative Neuroimaging

Structural and diffusion scans were acquired. All MR imaging was achieved using a GE Discovery MR750 3 Tesla device (GE Healthcare, Milwaukee, WI, U.S.A.) equipped with an 8-channel head coil.

### Diffusion Tensor Imaging

DTI was acquired with one image with a *b* value of 0 seconds/mm^2^ and 30 isotropically distributed diffusion directions with *b* values of 1000 seconds/mm^2^. Imaging parameters were as follows: TR/TE = 8000 milliseconds/60.7 milliseconds, slice thickness/increment = 2 mm/2 mm, flip angle = 90 degrees, matrix size = 128 × 128 millimeters, field of view = 256 millimeters. DTI processing was achieved using DSI Studio (http://www.dsi-studio.labsolver.org). The b-table was checked by an automatic quality control routine in DSI Studio to ensure its accuracy ^16^.

### Structural Imaging

For accuracy and comparison, fluid attenuation inversion recovery (FLAIR) sequences were overlaid onto the tensor or fODF map: Sagittal CUBE FLAIR sequences utilized a TR/TE: 6000 milliseconds/133 milliseconds, slice thickness/increment = 1.2 millimeters/0.6 millimeters, flip angle = 90 degrees, matrix = 256 × 256 millimeters, field of view 240 millimeters.

### Deep Learning Model

We adapted the deep learning model proposed by Nath et al. (2020). The input consists of 3 × 3 × 3 × 45 cubic patches of voxels. The cubic patch consists of 8th order diffusion-weighted magnetic resonance imaging spherical harmonic (SH) coefficients for each voxel in the patch (see *note on spherical harmonics*). The algorithm predicts the center voxel using spatial information as input features for the deep learning network. The loss function used was:

Where *m* denotes the number of samples, *y*_*true*_ is the set of fODF SH derived from MT-CSD, *y*_*pred*_ is the set of SH predictions made by ResCNN, *P*_*true*_ is the vector of tissue fraction value and *P*_*pred*_ is the predicted vector of tissue fractions. The ResCNN architecture is divided into three parts, its core part being the residual block which consists of multiple functional units, with each unit dedicated to a specific order of the SH. A 3D ResCNN architecture with a residual block was utilized. Hyper-parameters included a batch size of 1000, a learning rate of 1e^-4^, activation using ‘relu’, 40 training epochs with 250 steps per epoch and a validation frequency of 1. The DTI-datasets of the two clinical cases were then passed into the trained algorithm to yield MT-CSD approximations.

Data from 15 healthy subjects from the HCP were utilized for training and validation of the deep learning model in a training-validation-testing split of 5-2-8. The standard HCP acquisition protocol consists of 90 gradient directions for 3 b-values of 1000, 2000 and 3000 s/mm^2^. To ensure that this study would permit for processing of single-shell DTI data encountered in clinical settings, we utilized the diffusivity shell of 1000 s/mm^2^ with 90 gradient directions only for each subject, as this mimics the acquisition protocol of clinical DTI sequences. The diffusion weighted volumes of that specific shell were fitted to 8th order SH, which acted as input training data. The outputs were SH coefficients (8th order) of the fODF.

### Note on Spherical Harmonics

SH representations are a well-known, low-error representation of the DW-MRI signal ^17^ that remove the dependency of the number of gradient directions. Our model was trained with 8^th^ order SH representation, in particular training at higher orders allows for flexibility of utilizing lower SH orders by zero padding. The ResCNN architecture provides outputs of MT-CSD at 8^th^ order ^8^ along with voxel fractional volume estimates.

### Fiber Tractography

All tractography was performed by loading the tensor maps (for the DTI datasets) and fODF maps (for the post-processed deep learning algorithm datasets) into DSI Studio (http://dsi-studio.labsolver.org). Two experts in neuroanatomy and tractography (SP and JFM) performed the tract-seeding, cleaning and qualitative, and quantitative analytical portions of the study.

For the DTI data a variable fractional anisotropy (FA) parameter was utilized that permitted for maximal realistic tract representation, with minimal generation of spurious or false fibers. A deterministic streamline tracking approach ^12^ was utilized for generating tracts. Other tractography parameters included a differential tracking threshold of 0.2, an angular threshold of 0, a step size of 0.10 millimeters, a minimum fiber length of 30.0 millimeters, a maximum fiber length of 300.0 millimeters and automatic termination of tracking algorithm when 10,000 tracts were generated.

For the ODF-map outputs of the deep learning algorithm a quantitative anisotropy (QA) ^18^ threshold of 0.1 was utilized in each instance. This ensured that generation of spurious fiber tracts was minimized while preserving maximal anatomical detail. Other tracking parameters included a differential tracking threshold of 0.2, an angular threshold of 0, a step size of 0.10 millimeters, a minimum fiber length of 30.0 millimeters, a maximum length of 300.0 millimeters and termination of tracking algorithm when 10,000 tracts were generated.

In both control (DTI) and test (deep learning) cases, the region-of-interest (ROI)-based approaches were utilized to generate fiber tracts, based upon expert tractography knowledge, and also methods explained in our previous anatomical publications. Tracts generated included the arcuate fasciculus (AF) ^19^, superior longitudinal fasciculus (SLF) ^20^, inferior fronto-occipital fasciculus (IFOF) ^21^, uncinate fasciculus (UF) ^21^ and inferior longitudinal fasciculus (ILF) ^22^. For the corticospinal tracts (CST), we utilized a technique involving identifying vertically travelling fibers at the level of the pons using a color-coded diffusion map ^7^ and placing a seed ROI, together with another conduit-ROI within the vertically-oriented fibers at the level of the internal capsule. For the frontal aslant tract (FAT) ^23^ a seed-ROI was placed within the vertically-oriented sheet of fibers located within the dorsal aspect of the frontal lobe, together with a larger spherical ROI placed within the approximate level of the ventrolateral frontal lobe. For the cingulum, a single ball-shaped conduit-ROI was placed within the anterolaterally-travelling fibers within each hemispheric cingulate gyrus. Nevertheless, due to anatomical distortion produced by the tumors, variations in the ROI and seed placements were warranted, and guided by our anatomical expertise, in order to generate deviated or infiltrated tracts ^7^.

### Tract Volumes

Once tracts were generated we calculated their volume for comparison purposes using a technique utilized previously ^19–22^. We used the inbuilt function in DSI Studio, which calculates the number of voxels occupied by each fiber trajectory (streamlines) and the volume of the tracts (in milliliters) of each tract bundle. Volume of identical tracts (e.g. CST, AF, SLF etc.) in each hemisphere were compared between the DTI and deep learning methods, and the difference was calculated by subtracting the volume of the tracts generated using the deep learning method from the volume of the tracts generated using DTI.

## Results

### Diffusion Indices and Fiber Orientation Maps

When comparing diffusion maps, our results showed good consistency between the control (DTI) and test (deep learning) datasets. In subject 1, peritumoral areas with significant edema appeared highly isotropic, i.e. darker than the identical areas of the control FA maps **(Figure 1C, D)**. This phenomenon was also apparent when visualizing the directional color-coded FA and ODF maps which demonstrated substantial deviation of white matter caused by tumor mass effect **(Figure 1E, F)**. For subject 2, the tumor was smaller in comparison to subject 1, and with less corresponding edema. Nevertheless, the tumor area appeared more isotropic (darker) on the QA map versus the FA map **(Figure 2C, D)**. White matter architecture, as evidenced by directional color maps, was well-represented using both techniques, and despite the presence of space occupying lesions of varying size and associated edema **(Figure 2E, F)**.

### Qualitative Comparison

#### Subject 1

Due to the considerable peri-tumoral edema and midline shift, most of the right-sided white matter tracts were considerably deviated. Bilateral CSTs were visualized in both the DTI and deep learning datasets. Nevertheless, the post-processed CSTs were visually more robust on both sides versus the DTI CSTs. The right CST was deviated around the edematous area in the parietal lobe. Though DTI managed to replicate this deviation, the higher angular resolution of the algorithmic output portrayed a qualitatively superior visualization of the deviated CST fibers **(Figure 3A, B, C, D)**. Similarly, the right cingulum bundle was deviated towards the midline secondary to mass effect. This phenomenon was readily visualized in tracts using both methods, however the deep-learning technique produced bundles which were qualitatively larger-appearing, and occupying a greater volumetric area **(Figure 3E, F)**. Tumor edema extending to the ventrolateral frontal lobe caused the ventral aspect of the FAT to be pushed dorsally. Though this deviation was visualized using DTI, again the tracts generated using the deep learning dataset produced FAT bundles that were more robust **(Figure 4A, B)**. The right-sided AF and SLF were not found due to presence of tumor and edema. Both tracts were found on the left, with the deep learning dataset producing tracts that were thicker and occupying a larger proportion of white matter volume compared to those generated using DTI **(Figure 4C, D)**. The ILF was severely deviated secondary to edema extending into the temporal lobe, as demonstrated by both DTI and the deep learning dataset. The right-sided ILF of the latter nevertheless appeared more robust when compared to the DTI-generated ILF. Likewise, the mid-portion of the ILF in the external capsular area was pushed towards the midline, which was visualized using both techniques. Unlike other tracts, however, there was little qualitative difference between the IFOFs generated using either technique. Finally, the UFs were largely unaffected by the tumor or mass effect. Nevertheless, the UF was the only fiber bundle that was qualitatively superior in terms of bundle-thickness when generated using DTI **(Figure 4E, F)**.

**Figure 3.**
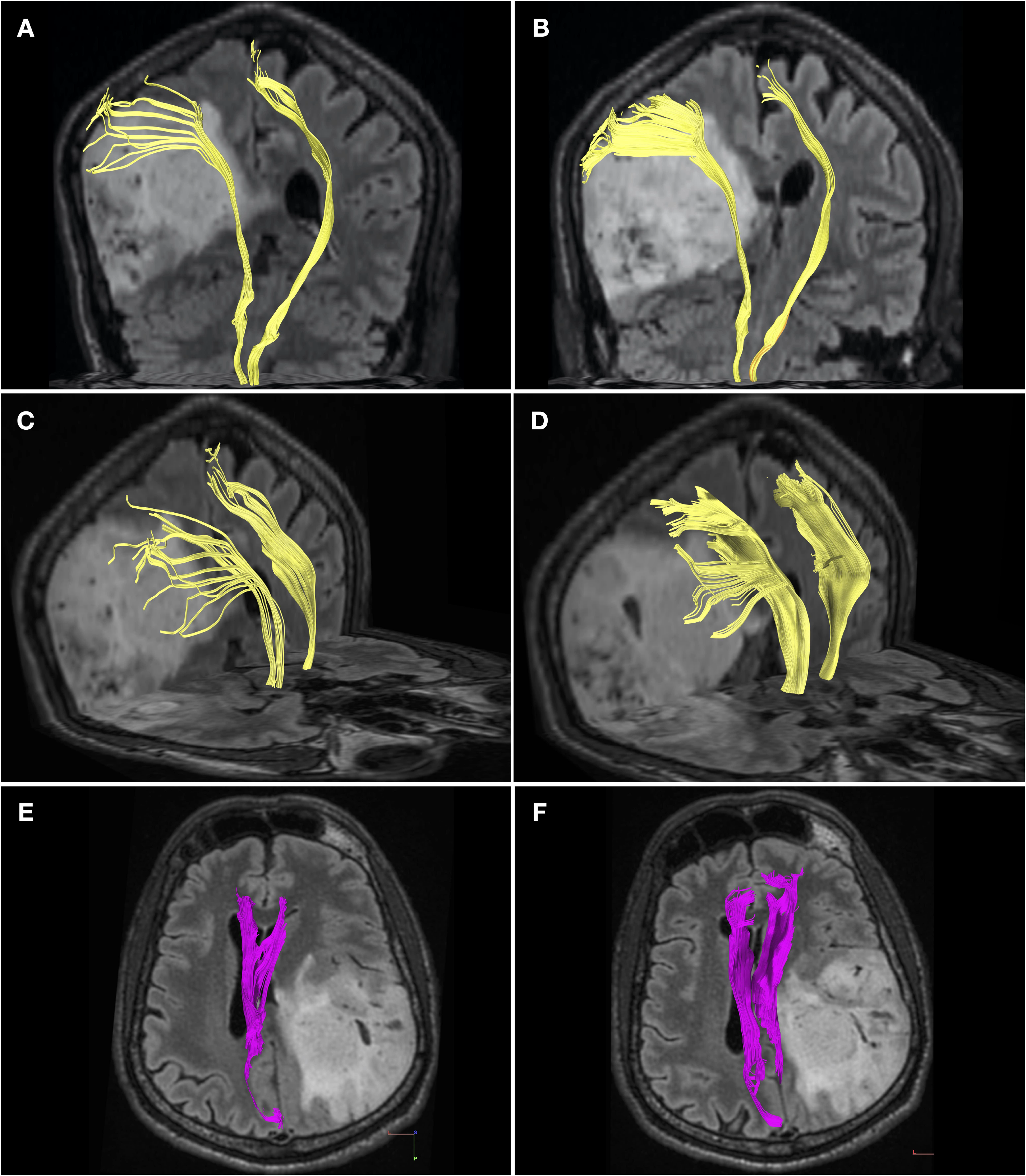
**A** – Coronal view of the DTI-generated CSTs in subject 1. Apparent is the degree of medial-deviation of the right CST produced by the tumor mass effect. **B –** Coronal view of the deep learning algorithm generated CSTs in subject 1. Compared to 3A, the tracts appear more robust, especially the laterally extending fibers of the corona radiata. **C –** An oblique-anterior view of the DTI CSTs in subject 1, here the shape of the tumor/edema has created an according deviation of the laterally-travelling CST fibers. **D -** An oblique-anterior view of the deep learning algorithm CSTs in subject 1. Compared to figure 3C, the tracts appear more robust along their entire course, which is especially apparent when comparing the laterally-travelling fibers of the corona radiata. **E –** Axial (supero-inferior) view of the DTI cingulum tracts in subject 1. Though the length of the left cingulum has been produced, only a portion of the right has, which is substantially deviated to the contralateral hemisphere. **F –** Axial (supero-inferior) view of the bilateral cingulum tracts created using the deep learning algorithm. Compared to 3E, not only has a much larger portion of the deviated right cingulum been reproduced, both bundles appear more robust.

**Figure 4.**
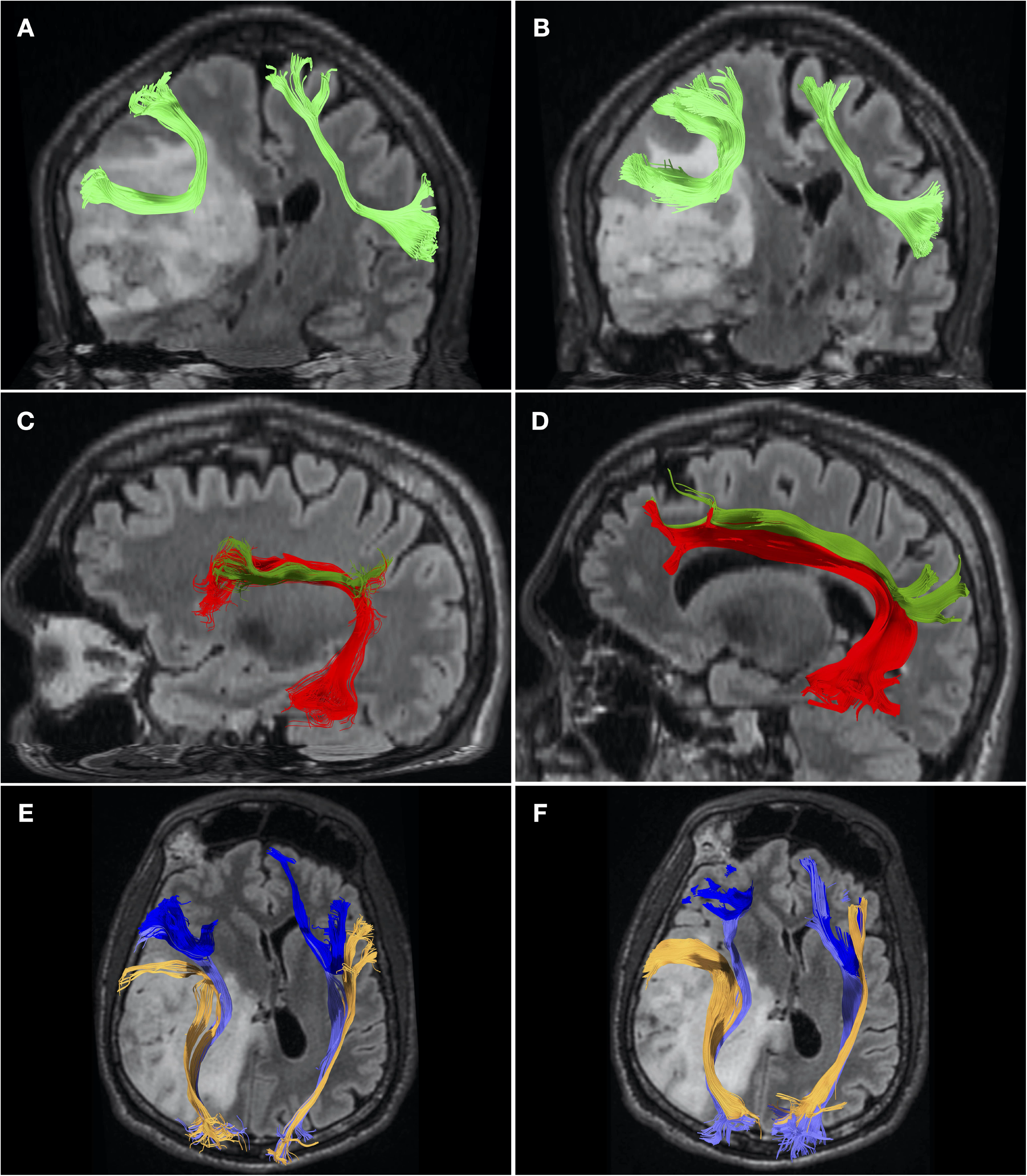
**A –** Coronal view of the bilateral DTI-generated FATs in subject 1. Apparent is the dorsal deviation of the ventral-component of the right FAT. **B –** Coronal view of the bilateral FATs created using the deep learning algorithm. Compared to 4A, the bundles appear more robust, with a greater number of recreated fibers. **C –** Sagittal view of the left sided AF (red) and SLF (green) bundles created using DTI. **D –** Sagittal view of the left sided AF (red) and SLF (green) bundles created using the deep learning algorithm. Compared to 4C, the tracts are both thicker and occupy a greater area of white matter volume. **E –** Axial (infero-superior) view of the IFOF (light purple), ILF (orange) and UF (blue) created using DTI. The right sided ILF and IFOF are severely deviated by mass effect. **F –** Axial (infero-superior) view of the IFOF (light purple), ILF (orange) and UF (blue) created using the deep learning algorithm. The ILF and IFOF on the tumor affected side are more robust compared to those in 4E. In particular, the right ILF demonstrates substantial deviation produced by edema extending into the temporal lobe.

#### Subject 2

In comparison to subject 1, subject 2 had a considerably smaller lesion with less peri-tumoral edema, and no significant mass-effect. The CSTs were readily visualized using both DTI and the deep learning technique. Nevertheless, CSTs generated using the latter method were more robust in terms of bundle size, and possessed the laterally-extending fibers of the corona radiata which were absent in the DTI tracts **(Figure 5A, B, C, D)**. Bilateral cingulum bundles were readily depicted using both DTI and the deep learning method. Again, the latter method resulted in qualitatively more robust bundles **(Figure 5E, F)**. The biggest qualitative difference was observed in the frontal aslant tracts, which appeared markedly larger using the deep learning method **(Figure 6A, B, C, D)**. Unlike subject 1, the smaller amount of peri-tumoral edema did not preclude visualization of the right-sided AF and SLF **(Figure 6E, F)**, which again were better depicted using the deep learning technique. This trend was again preserved for the ventral fronto-temporal tracts, with the exception of the UF, which like in subject 1, was markedly larger when visualized using DTI **(Figure 7A, B, C, D, E, F)**.

**Figure 5.**
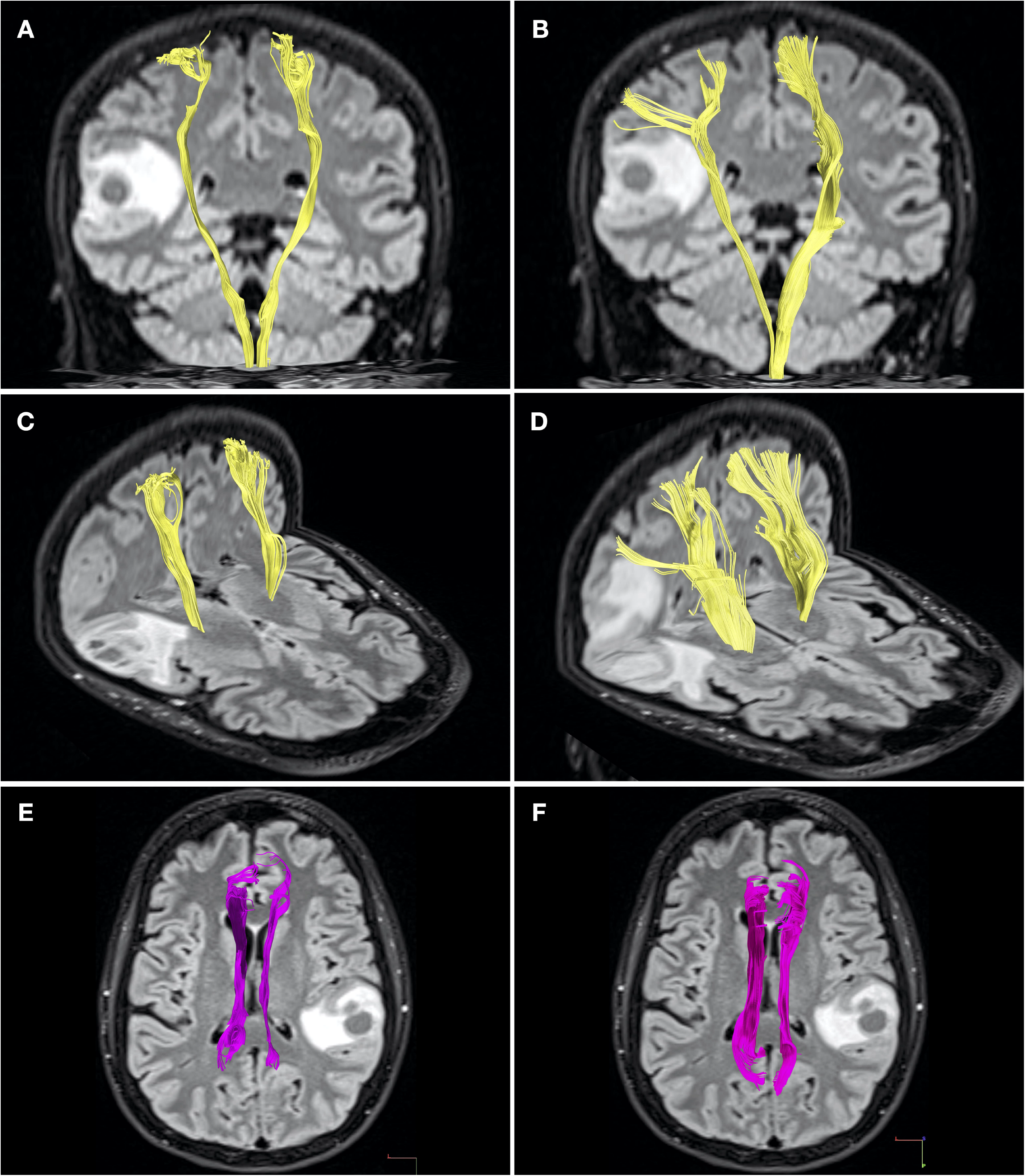
**A –** Coronal view of the DTI-generated CSTs in subject 2. There is slight thinning of right-sided CST fibers when compared to the contralateral hemisphere. **B –** Coronal view of the CSTs generated from the deep learning dataset, overall, both CSTs are more robust when compared to the DTI CSTs (5A). Moreover, laterally-extending fibers of the corona radiata are reproduced, which are absent on the DTI CSTs. **C –** Oblique-anterior view of the DTI-generated CSTs in subject 2. **D –** Oblique-anterior view of the CSTs generated using the deep learning dataset. Not only are the CSTs more robust versus those in 5C, the laterally-extending fibers of the right corona radiata are readily visualized. **E –** Supero-inferior axial view of the bilateral cingulum bundles generated using DTI. The right cingulum is thinned versus the left, despite not being directly impinged upon by tumor or edema. **F –** Supero-inferior axial view of the bilateral cingulum bundles generated using the deep learning dataset. Compared to 5E, both bundles are qualitatively thicker in general, but there is still some apparent thinning of the right-sided cingulum.

**Figure 6.**
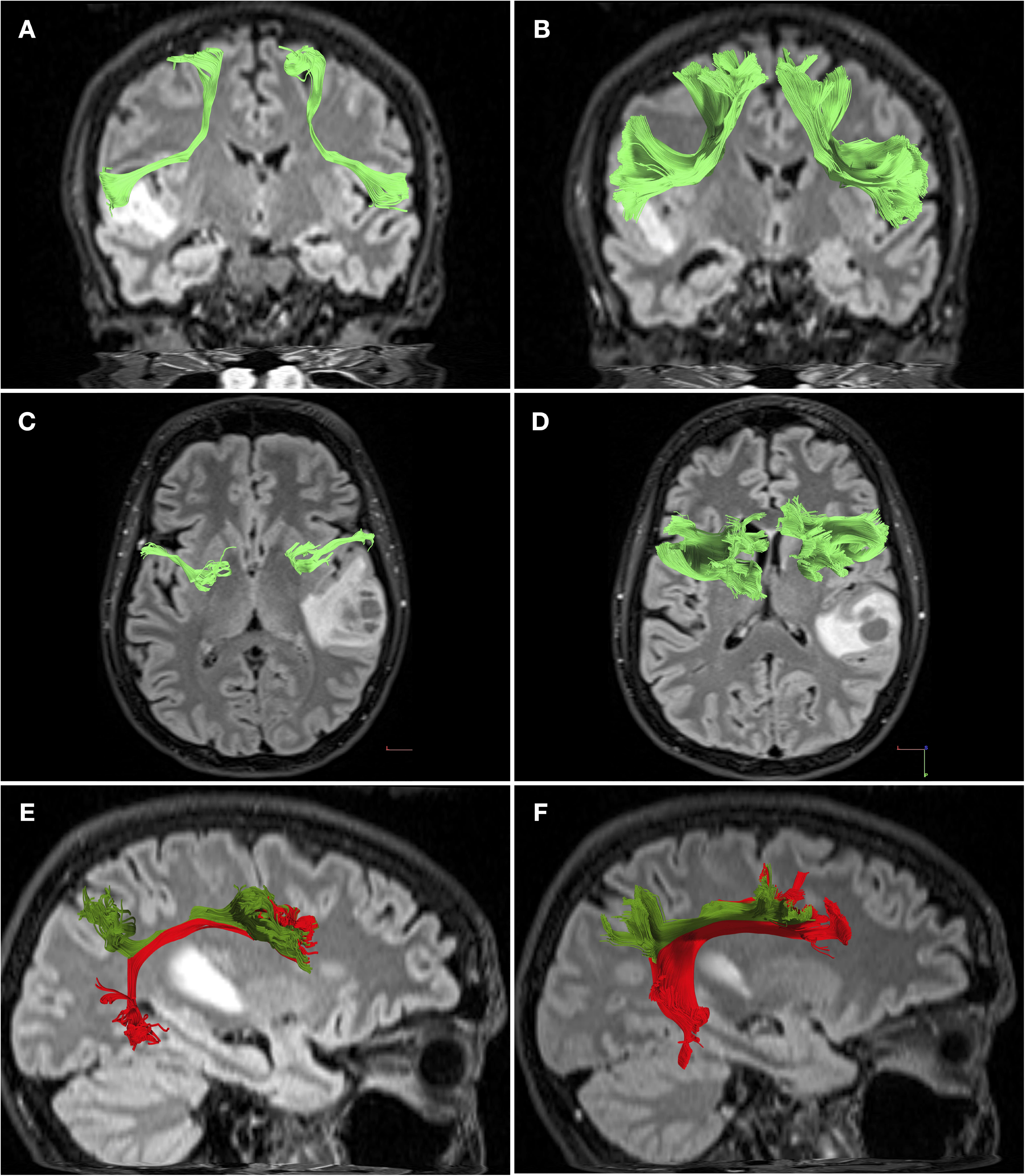
**A –** Antero-posterior coronal view of subject 2’s bilateral FATs generated using DTI. There is no apparent deviation of the right-sided FAT. **B –** Antero-posterior coronal view of the bilateral FATs generated using the deep learning dataset. There is a striking difference between the FATs generated using this method, compared to the DTI-generated FATs (6A). **C –** Supero-inferior axial view of subject 2’s bilateral FATs generated using DTI. Apparent is how thin the generated bundles are, bilaterally. **D –** Supero-inferior axial view of subject 2’s bilateral FATs generated using the deep learning algorithm. These are substantially larger in volume and bundle thickness versus those generated using DTI (6C). **E –** Right-sided sagittal view of the AF (red) and SLF (green) generated using DTI in subject 2. **F –** Right-sided sagittal view of the AF (red) and SLF (green) generated using the deep learning dataset in subject 2. Compared with 6E, both tracts are more robust.

**Figure 7.**
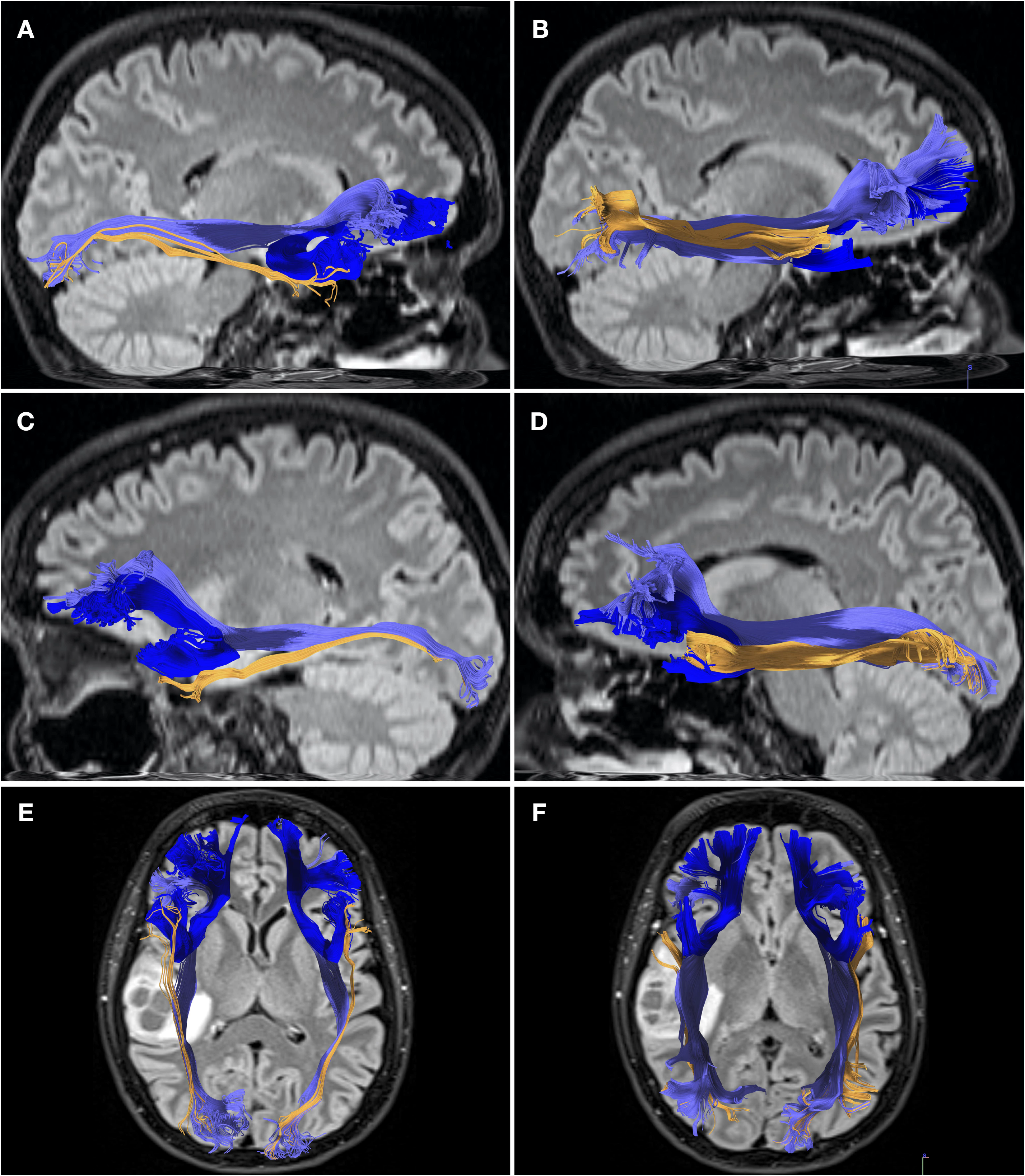
**A –** Right-sided sagittal view of the DTI-generated IFOF (light purple), ILF (orange) and UF (blue), in subject 2. **B –** Right-sided sagittal view of the IFOF (light purple), ILF (orange) and UF (blue), generated using the deep learning dataset in subject 2. In particular, the IFOFs medial frontal terminations are present, while they are absent on the DTI generated IFOF (7A). The ILF is also more robust compared to the DTI-generated ILF. **C –** Left-sided sagittal view of the DTI-generated IFOF (light purple), ILF (orange) and UF (blue), in subject 2. **D –** Left-sided sagittal view of the IFOF (light purple), ILF (orange) and UF (blue), generated using the deep learning dataset in subject 2. The IFOF and ILF are both markedly larger than those created using the DTI dataset (compare with 7C). **E –** Infero-superior axial view of the DTI-generated IFOF (light purple), ILF (orange) and UF (blue), in subject 2. Here, the apparent difference in tract sizes of both ILF and IFOF are pronounced, compared to the DTI-generated tracts demonstrated in 7E.

### Quantitative Comparison

#### Subject 1

Tract bundles generated using data processed by the deep learning algorithm were in general quantitatively larger in volume when compared to tracts created using DTI. The only tracts that were larger in volume when generated using the DTI dataset were the UFs **(Table 1)**.

#### Subject 2

Tract bundles generated using data processed by the deep learning algorithm were in general quantitatively larger in volume when compared to tracts created using DTI. The only tracts that were larger in volume when generated using the DTI dataset were the UFs **(Table 1)**.

## Discussion

We successfully employed a deep learning algorithm to transform clinically-acquired DTI data into non-tensor high-angular resolution data, permitting tractography, in two glioma patients. All tracts found within the DTI dataset were also replicated within the deep learning dataset, and the deep learning method produced qualitatively superior and quantitatively larger tract bundles.

### Clinical Considerations

Of the numerous adjunctive techniques available to surgeons, pre-surgical tractographic white matter visualization is advantageous as it can be achieved non-invasively and pre-operatively. Consequently, accurate depiction of white matter tracts is important, especially those in close proximity to the tumor or passing within the plane of approach. Tensor-based techniques have been shown to be less reliable than non-tensor techniques at reproducing fiber tracts passing through, or near to aberrant or edematous structures ^1,5–7,24–26^. Peri-tumoral DTI tracts may therefore appear to terminate abruptly, be thinned, or be entirely absent^5,7^. This phenomenon is evident in this study, when comparing tracts generated using DTI with those using the post-processed deep learning datasets.

Using our technique, any institution possessing a DTI-capable MR device can theoretically obtain high-resolution tractography without acquisition of research-grade hardware or personnel. Furthermore, scanning time may be substantially reduced making high-quality tractographic reconstructions clinically feasible: For example, a single shell-shell, 32 direction scan may take approximately 5 minutes to acquire, whereas a 3-shell, 90 direction scan may take approximately 40 minutes^14^. Our approach demonstrates that high-quality tractography can be conducted in short, clinically-feasible timeframes.

By dispensing with specialized hardware or scanning protocols, our approach may also alleviate some of the costs associated with obtaining high-quality white matter imaging. It does, however, necessitate the expertise of personnel who are sufficiently proficient in computer programming languages, with technical knowledge of machine learning/deep learning and radiological data formats (i.e. digital communication and imaging in medicine (DICOM)).

### Technical Considerations and Limitations

Despite the general quantitative and qualitative improvement in white matter tract reproduction produced by the deep learning algorithm, neither DTI nor the algorithm could reproduce tracts (namely the AF and SLF) within the highly edematous, peritumoral region of subject 1. This appears to be a general shortcoming of all tractography approaches, and it highlights the potential for complimentary adjunctive techniques, such as intraoperative cortical stimulation^2^. For example, the tracts may still be intact but unable to be visualized tractographically, due to presence of edema, tumor or necrosis ^5,7^.

A further technical consideration remaining to be accounted for is that DTI results in qualitatively and quantitatively superior reproduction of the UFs, whereas the deep learning algorithm was better at reproducing all other tracts. We postulate that this could be secondary to the relatively short length of UF fibers, its acute turning angle over a small area, and the fact that the UF shares terminations with other ventral tracts like the IFOF and ILF.

## Conclusions

In our study, we have successfully demonstrated qualitatively and quantitatively superior white matter tractography results produced by a novel deep learning algorithm applied to standard DTI data. This method, which yields higher quality tractography provides a more complete anatomical picture. It may be particularly useful to neurosurgeons for pre-operative planning.

## Data Availability

All data used in this paper can be obtained by contacting the relevant authors.

## Notes

### Competing Interest Statement

The authors have declared no competing interest.

### Funding Statement

No funding received.

